# The Inflammatory Cascade Through Discrimination, Socio-economic Status, and Body-Mass Index

**DOI:** 10.64898/2026.06.24.26356254

**Authors:** Michael J. Espero

**Affiliations:** Claremont Graduate University

## Abstract

C-Reactive Protein (hs-CRP) is a common marker for human inflammation, a response to perceived threat and precipitate to many compromising health conditions. Previous work demonstrated that in addition to other biological features that may be predictive and explanatory of variance in inflammation, psychosocial influences may play a role. The present work uses structural equation modeling to examine pathways including socioeconomic status (SES), psychological capital (PsyCap), and perceived discrimination (Discrim)-insofar as they explain variance in hs-CRP, potentially moderated by neurological lateralization (handedness). Body mass index (BMI), an indicator of body composition, stood as the strongest predictor of the obesity-related inflammatory marker (ORIM). On average, females are predicted to have higher hs-CRP scores than males. The psychosocial constructs were estimated to have little to no effect on inflammation (via hs-CRP) in the analysis sample (ADD Health Study) in either group (left and right-handers) although a small, statistically non-zero indirect path is found in the retained model for right-handed participants (given statistical power for estimation). With this finding, contextual effect estimates are provided with regard to the effect of perceived discrimination on hs-CRP given the range of SES and BMI.

## Inflammation and Health

The general consensus of the purpose of the inflammatory response is to restore one to optimal homeostatic state when faced with injury or other challenging situations (Antonelli and Kushner, 2019; Sorrells and Sapolsky, 2007). While acute inflammation can be seen as a means to restoration (stimulation), ongoing, chronic inflammation is thought to be associated with poorer (relative to periods before the physical or psychosocial offense) digestive, immune, reproductive, and cognitive function. Ridker, Hennekens, Buring, and Rifai (2000) found C-Reactive Protein (hs-CRP), a marker of inflammatory response, to be the strongest predictor of cardiovascular events in a sample (n = 28,263) of healthy, post-menopausal women. Those whose measurements of hs-CRP that stood in the highest quartile (compared to the lowest) were estimated at 4.4 times increased relative risk (95% CI: 2.2-8.9) in a univariate model and 1.5 (95% CI: 1.1-2.2) in a multivariate model. Prediction of gum disease (periodontitis), inflammatory conditions that affect tissues surrounding the teeth, was bolstered by analysis of variance including hs-CRP as an independent variable when comparing subjects with deep periodontal pockets (spaces that form between the gums and teeth) with shallower pockets, *p* < .001, (Pejcic, Kesic and Milasin, 2010). Moreover, Tamakoshi, Yatsuya, Kondo, Hori, Ishikawa, Zhang, Murata, Otsuka, Zhu, and Toyoshima (2003) found links between hs-CRP and metabolic syndrome (MS), identifying a continuum of MS components (hypertension, impaired glucose tolerance, hyper-insulinemia, increased levels of triglyceride, decreased level of HDL cholesterol, and obesity) to follow with increased likelihood of elevated hs-CRP with the addition of each MS component (1-5 components), 1.48 (95% CI: 1.21–1.81), 1.84 (95% CI: 1.51–2.24), 1.91 (95% CI: 1.54–2.39), 3.42 (95% CI: 2.66–4.41), 4.17 (95% CI:3.21–5.41). The inflammatory marker, hs-CRP, fits as a substantial element in efforts to explain and predict health outcomes.

## Discrimination

The notion that perceived social discrimination can negatively impact human health is not new. In 2010, work from Lewis, Aiello, Leurgans, Kelly, and Barnes indicated a statistically non-zero, small, positive association between everyday discrimination and hs-CRP (n = 296; Minority Aging Research Study), *β* = 0.11, *p* = 0.02. This finding, however, was challenged and attenuated when BMI was added as a covariate, *β* = 0.09, *p* = 0.07. K. Sims, M. Sims, Glover, Smit, and Odden (2020) found increased inflammatory response as indexed by hs-CRP for African Americans (n = 5,145; Jackson Heart Study) when groups that reported never experiencing everyday discrimination were compared with those who experienced *more* frequent discrimination, adjusting for clinical (i.e. HbA1c, HDL), behavioral (i.e. physical activity, smoking), and socioeconomic characteristics (i.e. education level, income group), *β* = 0.07, (95% CI: 0.1 - 0.12).

## Socio-Economic Status

While much of the world has experienced improvement in overall health status and life expectancy when considering longer time-scales, for most nations, those of relatively lower socio-economic status (SES) faced higher mortality rates (Feinstien, 1993). SES, defined as a measure of a person’s social and economic standing relative to others, stands as an important construct with potential to influence systemic inflammation and possibly moderate the effects of discrimination on inflammatory response among other important health outcomes. Liu, Aiello, Mensah, Gasser, Rueb, Cordel, Juonala, Wake, and Burgner (2018) meta-analyzed 21 studies (n = 43,629), finding an overall 25% increased average hs-CRP when comparing those grouped in the lowest childhood SES to those in the highest, ratio change in hs-CRP = 1.25, (95% CI: 1.19 - 1.32). Critically, when BMI was added to their model, the association between childhood SES and adult hs-CRP was *fully* attenuated, ratio change in hs-CRP = 1.04, (95% CI: 0.93 - 1.13). Differences in education (years) served as a predictor of hs-CRP for Stepanikov, Bateman, and Oates (2017). After including adjustment for perceived daily and lifetime discrimination, education (years) stood as a statistically non-zero predictor (*β* = -0.04, *p* < 0.01), although substantially less influential than race (Black), which was estimated to influence relatively higher inflammatory response (compared to other groups), *β* = 0.36, *p* < 0.01. However, when psychological factors (i.e. depression, anxiety), lifestyle factors (i.e. smoking, physical activity), **BMI**, and other health factors (i.e. hypertension, diabetes) were added to the model, the estimated influence of both race and education was attenuated and the statistically non-zero estimated effects that remained (predicting hs-CRP) were diabetes (*β* = 0.31, *p* < 0.01; increasing hs-CRP, on average) and hypertension (*β* = -0.24, *p* < 0.01; on average, associated with relatively *decreased* hs-CRP), and BMI, *β* = 0.01, *p* < 0.001.

## Psychological Capital

Distinguished from what one *has* (economic capital), what one *knows* (human capital), and *who* one knows (social capital) - psychological capital (PsyCap) refers to *who you are* in the sense of one’s capacities of confidence, hope, optimism, and resilience (Luthans, Luthans, and Luthans 2004). With regard to explaining variance in a hs-CRP (Log-CRP) outcome in a sample of 944 middle-aged Swedes, Marteinsdottir, Ernerudh, Jonasson, Kristenson, and Garvin (2016) modeled dimensions of psychological resources (theoretically oblique to dimensions of PsyCap), estimating statistically non-zero effects for self-esteem (protective; *β* = -0.11, *p* = 0.021), hopelessness (harmful; *β* = 0.14, *p* = 0.014), depression (harmful; *β* = 0.12, *p* = 0.012), but not coping (*β* = -0.08, *p* = 0.078)-after adjusting for age and sex. However, when the covariates smoking, alcohol intake, fruit & vegetable intake, physical activity, body mass index, blood pressure, blood lipids, and blood glucose were added to the modeling process, only the protective estimate for the influence of self-esteem remained, *β* = -0.09, *p* = 0.037. Although the effects of psychological capital/resources were largely diminished in the authors’ full model, it is worth noting that another inflammatory marker, IL-6, modeled with the full covariate set-yielded non-zero estimates for each PsyCap-related dimension sampled (except hopelessness).

## Handedness in Psychological Research

While research methodologists working in fields of psychology and neuroscience may take heed of approaches to recruit samples of people representative of the populations they wish to generalize to, considering demographic facets such as sex, gender, ethnicity, educational background, and income - similar efforts based on population-level generalizability for handedness (left and right-dominant manual tool use) may be less defensible throughout decades of research - to the extent that the wealth of psychology and neuroscientific study amounts to largely the study of right-handed people (more so than population prevalence would suggest). Bailey, McMillan and Newman (2019) found that given 1,008 peer-reviewed papers published in journals such as *Cerebral Cortex*, *Human Brain Mapping*, and *Neuroimage* with data from 86,640 human subjects, handedness data was available for 31,973 - of whom 3.22% (1,031) were reported left-handed or ambidextrous. Synthesis of meta-analyses (n = 2M+) by Papadatou, Ntolka, Schmitz, Martin, Munafo, Ocklenburg, and Paracchini (2020) estimate the population prevalence of left-handedness to be 10.4% (∼ 1 in 10 people) - roughly triple the proportion observed by Bailey et al. (2019) across a large sample of neuropsychological publications. As a unobtrusive indicator (albeit imperfect) of atypical cerebral organization and language lateralization (Giovagnoli & Parisi, 2024), handedness is included in the present work; this is intended to allow the comparisons of path estimates and the relations of psychosocial constructs to the manifest indicator of inflammation, hs-CRP.

## Hypotheses

1. Discrimination → BMI → hs-CRP. The effects of felt discrimination on hs-CRP is partially mediated through BMI.
2. Discrimination → SES → BMI → hs-CRP. The effects of felt discrimination on hs-CRP is mediated through SES and BMI. Discrimination may negatively influence SES, which may, in turn, increase BMI, leading to higher hs-CRP.
3. Discrimination → PosPsyCap → BMI → hs-CRP. The effects of felt discrimination on hs-CRP is mediated through PosPsyCap and BMI. Discrimination may negatively influence PosPsyCap, which may, in turn, increase BMI, leading to higher hs-CRP.

## Methods

### Participants

The study included 1,522 (1,376 right-handers and 146 left-handers) participants (Figure 1. Participants) from the fifth wave of the National Longitudinal Study of Adolescent to Adult Health (ADD Health Study). The ADD Health Team emphasizes strict confidentiality procedures and ensures identifying information is not disclosed. Moreover, parental permission and informed consent were integral parts of the recruitment process drawing participants from many sites. An additional modification to the analysis data set included filtering to observations with hs-CRP (Figure 2. Distribution of hs-CRP by Handedness) less than or equal to 20.

**Figure 1.**
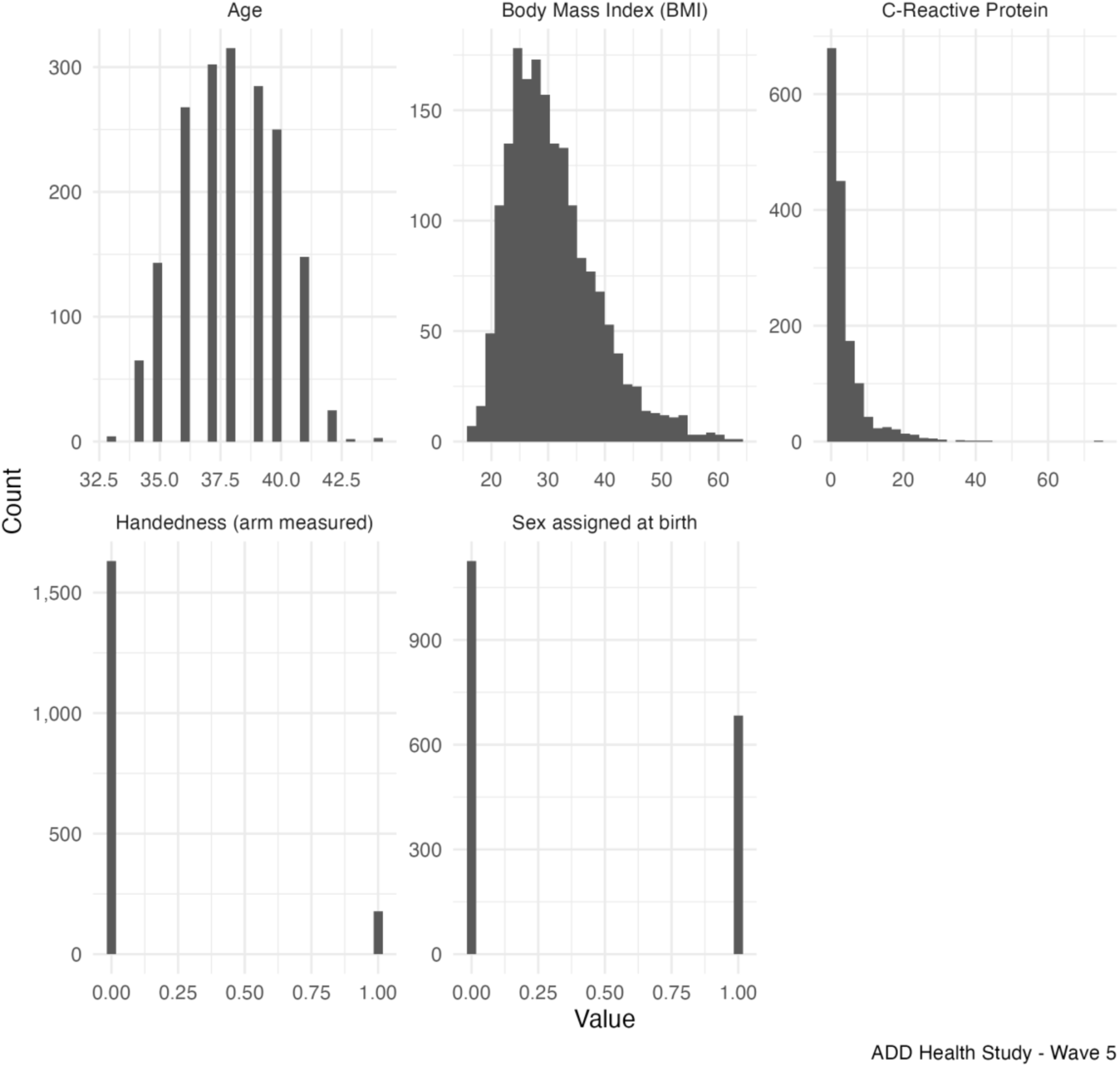
Participants

**Figure 2.**
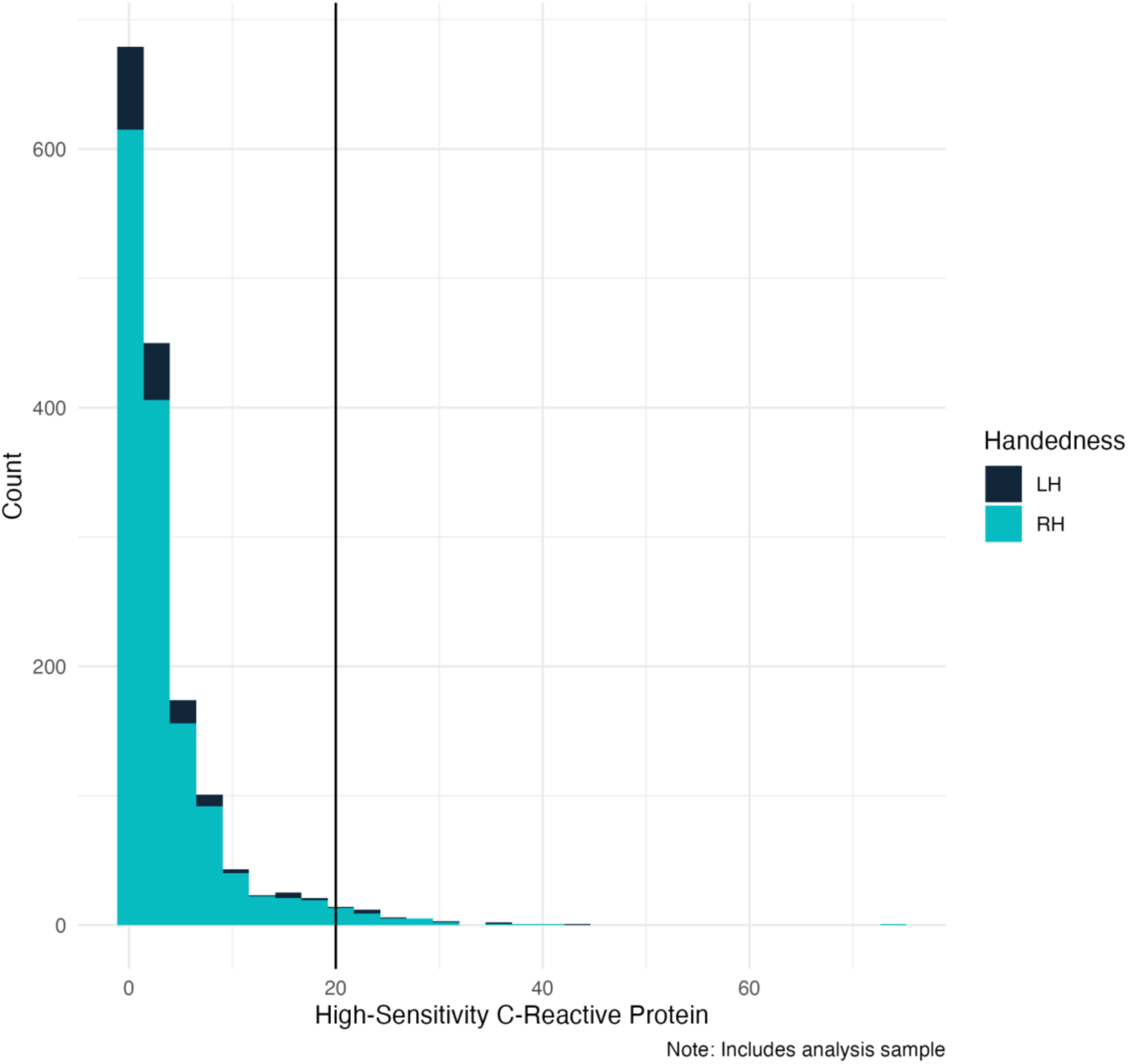
Distribution of hs-CRP (outcome) by Handedness ADD-Health Study Wave 5

### Analyses

Models were estimated in lavaan 0.6-19 with full information maximum-likelihood and Huber–White cluster-robust standard errors. Data were split by handedness—Right-handed (RH) and Left-handed (LH)—while clustering handled the nested school structure. Model fit was judged “adequate” if robust CFI greater-than or equal to .90; “good” if RMSEA less-than .06 and SRMR less-than .08. Distribution-free indirect effects, standard errors, and 95% confidence intervals were computed with the bootstrap method and two-tailed z-tests. Direct and indirect effects were deemed statistically significant if confidence intervals did not contain zero. Morover, Bayesian structural equation modeling is also provided, yielding distributional estimates of effects.

## Results

### SEM

#### Model Fit

Robust Chi-Squared (214) = 533.626, *p* less than .001 (sensitive to sample size); CFI =.927; TLI =.907; RMSEA = .05; SRMR =.045. Fit (Table 1. Model Fit Indices) met RMSEA/SRMR benchmarks; incremental indices are *acceptable*.

**Table 1.** Model Fit Indices.

| Table 1. Model Fit Indices |  |  |  |  |  |
| --- | --- | --- | --- | --- | --- |
| CFI | TLI | RMSEA | SRMR | AGFI | IFI |
| 0.927 | 0.907 | 0.05 | 0.045 | 0.991 | 0.927 |
| Note: CFI robust (Brosseau-Liard & Savalei, 2014) reported as CFI. |  |  |  |  |  |

#### Measurement Model

**Table 2.** Measurement Model.

| Table 2. Measurement Model |  |  |  |  |  |  |  |
| --- | --- | --- | --- | --- | --- | --- | --- |
|  |  | Right-Handers |  |  | Left-Handers |  |  |
|  | Indicator | Estimate | Std. Error | P-Value | Estimate | Std. Error | P-Value |
| SES | What is the highest level of education that you have achieved to date? | 2.029 | 0.104 | < .001 | 1.774 | 0.318 | < .001 |
|  | In the last calendar year, how much income did you receive from personal earnings before taxes? Include wages or salaries, tips, bonuses, overtime pay, and income from self-employment. | 1.784 | 0.110 | < .001 | 1.776 | 0.404 | < .001 |
|  | What was the total household income before taxes and deductions in the last calendar year for all household members who contribute to household expenses? | 1.917 | 0.083 | < .001 | 1.782 | 0.325 | < .001 |
|  | Pick the number for the step (hierarchical ladder analogy) that shows where you think you stand at this time in your life, relative to other people in the United States. | 1.280 | 0.069 | < .001 | 1.093 | 0.300 | < .001 |
| PosPsyCap | I am always optimistic about my future. | 0.306 | 0.029 | < .001 | 0.308 | 0.111 | 0.006 |
|  | I finish whatever I begin. | 0.515 | 0.031 | < .001 | 0.485 | 0.103 | < .001 |
|  | I am a hard worker. I keep working when others stop to take a break. | 0.468 | 0.033 | < .001 | 0.448 | 0.079 | < .001 |
|  | I am diligent. I never give up. | 0.617 | 0.027 | < .001 | 0.633 | 0.097 | < .001 |
| Discrim | You are treated with less courtesy or respect than other people. | 0.584 | 0.025 | < .001 | 0.733 | 0.080 | < .001 |
|  | You receive poorer service than other people at restaurants or stores | 0.491 | 0.023 | < .001 | 0.295 | 0.068 | < .001 |
|  | People act as if they think you are not smart. | 0.565 | 0.028 | < .001 | 0.471 | 0.063 | < .001 |
|  | People act as if they are afraid of you. | 0.324 | 0.027 | < .001 | 0.234 | 0.076 | 0.002 |
|  | You are threatened or harassed | 0.304 | 0.022 | < .001 | 0.308 | 0.057 | < .001 |
|  | I am always optimistic about my future. | 0.164 | 0.030 | < .001 | 0.217 | 0.093 | 0.020 |

Notes: 1. Clusters defined by 'CLUSTER2'. 2. Groups defined as right-handed (RH) and left-handed (LH). 3. The PosPsyCap indicator 'I am always optimistic about my future' was specified to cross-load on the Discrimination factor based on theoretical expectations that chronic experiences of discrimination systematically diminish one's outlook on the future.

#### Factor Reliabilities

**Table 3.** Factor Score Reliabilities.

| Table 3. Factor Score Reliabilities |  |  |  |
| --- | --- | --- | --- |
|  | SES | PosPsyCap | Discrim |
| RH | 0.76 | 0.77 | 0.78 |
| LH | 0.73 | 0.79 | 0.83 |
| These are the square of the factor determinacies, which represent the estimated correlation between factor scores and latent variable scores. |  |  |  |

### Structural Model

#### Direct Effects

**Table 5:** Parameter Estimates.

| Left-Handers |  |  |  |  |  |  | Right-Handers |  |  |  |  |  |  |  |  |
| --- | --- | --- | --- | --- | --- | --- | --- | --- | --- | --- | --- | --- | --- | --- | --- |
| Parameter | Estimate | Std. Error | $\beta$ | Z | P-Value | Lower | Upper | Parameter | Estimate | Std. Error | $\beta$ | Z | P-Value | Lower | Upper |
| SES ~ Discrim | -0.243 | 0.129 | -0.242 | -1.877 | 0.061 | -0.496 | 0.011 | SES ~ Discrim | -0.297 | 0.049 | -0.285 | -6.010 | < 0.001*** | -0.394 | -0.200 |
| PosPsyCap ~ Discrim | -0.047 | 0.149 | -0.043 | -0.312 | 0.755 | -0.340 | 0.246 | PosPsyCap ~ Discrim | 0.087 | 0.038 | 0.087 | 2.283 | < 0.05* | 0.012 | 0.162 |
| BMI ~ SES | -2.978 | 1.137 | -0.335 | -2.619 | < 0.01** | -5.208 | -0.749 | BMI ~ SES | -1.666 | 0.310 | -0.243 | -5.373 | < 0.001*** | -2.273 | -1.058 |
| BMI ~ PosPsyCap | 0.862 | 0.862 | 0.105 | 1.000 | 0.317 | -0.828 | 2.551 | BMI ~ PosPsyCap | -0.225 | 0.243 | -0.032 | -0.926 | 0.354 | -0.700 | 0.251 |
| BMI ~ Discrim | -0.312 | 0.897 | -0.035 | -0.348 | 0.728 | -2.070 | 1.446 | BMI ~ Discrim | 0.060 | 0.278 | 0.008 | 0.214 | 0.83 | -0.486 | 0.605 |
| BMI ~ SEX | -0.203 | 0.530 | -0.012 | -0.384 | 0.701 | -1.241 | 0.835 | BMI ~ SEX | -0.486 | 0.400 | -0.033 | -1.215 | 0.224 | -1.269 | 0.298 |
| CRP ~ BMI | 0.211 | 0.058 | 0.446 | 3.660 | < 0.001*** | 0.098 | 0.324 | CRP ~ BMI | 0.236 | 0.018 | 0.443 | 13.339 | < 0.001*** | 0.201 | 0.271 |
| CRP ~ SES | 0.069 | 0.386 | 0.016 | 0.178 | 0.859 | -0.687 | 0.825 | CRP ~ SES | -0.160 | 0.128 | -0.044 | -1.250 | 0.211 | -0.411 | 0.091 |
| CRP ~ PosPsyCap | -0.281 | 0.362 | -0.072 | -0.778 | 0.437 | -0.990 | 0.427 | CRP ~ PosPsyCap | 0.081 | 0.121 | 0.021 | 0.670 | 0.503 | -0.156 | 0.318 |
| CRP ~ Discrim | -0.205 | 0.525 | -0.049 | -0.391 | 0.696 | -1.235 | 0.825 | CRP ~ Discrim | 0.043 | 0.129 | 0.011 | 0.335 | 0.737 | -0.209 | 0.295 |
| CRP ~ AGE | -0.086 | 0.070 | -0.041 | -1.229 | 0.219 | -0.222 | 0.051 | CRP ~ AGE | -0.086 | 0.048 | -0.045 | -1.807 | 0.071 | -0.179 | 0.007 |
| CRP ~ SEX | 1.662 | 0.695 | 0.203 | 2.393 | < 0.05* | 0.301 | 3.024 | CRP ~ SEX | 1.346 | 0.168 | 0.173 | 8.020 | < 0.001*** | 1.017 | 1.675 |
| n = 146; clusters = 67 |  |  |  |  |  |  | n = 1,376; clusters = 130 |  |  |  |  |  |  |  |  |

#### Direct Effects. Left-handed Participants

Table 5 (above) displays that for left-handed participants (n = 146), the association between BMI and CRP was positive and statistically non-zero, *β* = 0.211, SE = 0.058, 95% CI (0.098 - 0.324). The association between SES and hs-CRP was estimated statistically no different from zero, *β* = 0.069, SE = 0.386, 95% CI (-0.687 - 0.825). With regard to sex, the association with the inflammation outcome is positive (females relative to males) and statistically non-zero, *β* = 1.662, SE = 0.695, 95% CI (0.301 - 3.024). For left-handers, the relationship between PosPsyCap and discrimination was estimated as statistically no different from zero, *β* = -0.047, SE = 0.149, 95% CI (-0.340 - 0.246).

#### Direct Effects. Right-handed Participants

Table 5 (above) displays that for right-handed participants (n = 1,376), the association between BMI and hs-CRP was positive and statistically non-zero, *β* = 0.236, SE = 0.018, 95% CI (0.201 - 0.271). The association between SES and hs-CRP was estimated as statistically no different from zero, *β* = -0.160, SE = 0.128, 95% CI (-0.411 - 0.091). With regard to sex, the association with the inflammation outcome is positive (females relative to males) and statistically non-zero, *β* = 1.346, SE = 0.168, 95% CI (1.017 - 1.675). With regard to psychological capital, discrimination, and age, direct effects were estimated to be no different than zero (consider this in the context of indirect effects estimated, Table 6, below). In this sample, the relationship between PosPsyCap and discrimination was estimated as non-zero for right-handers, *β* = 0.087, SE = 0.038, 95% CI (0.012 - 0.162).

A multigroup path diagram illustrating the hypothesized model is displayed below (Figure 3. Path Diagram).

**Figure 3.**
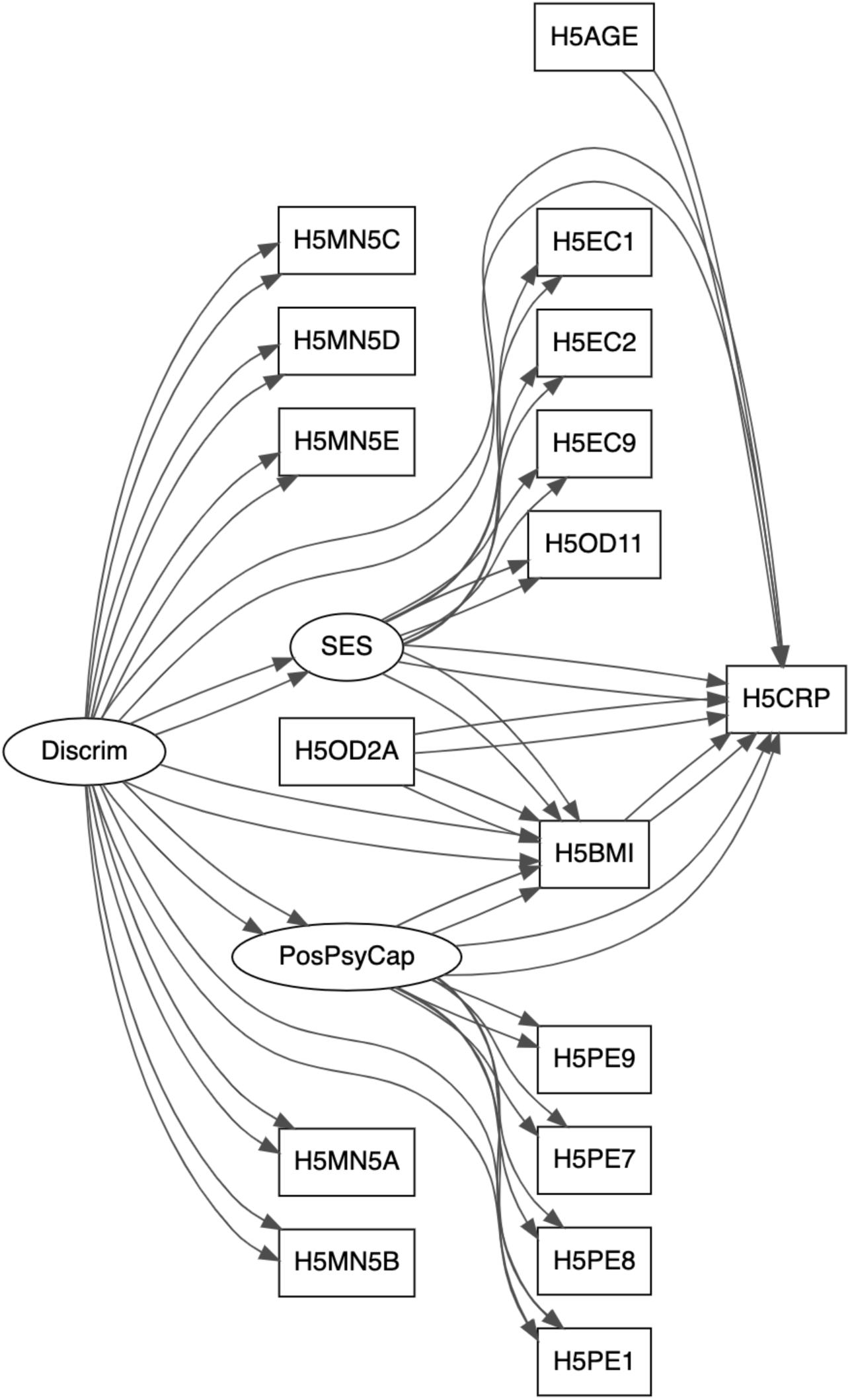
Path Diagram

#### Indirect Effects

The indirect effect (see Table 6. Indirect Effects, above; Figure 4 displays bootstrap estimates of the main indirect effect estimate) of perceived discrimination on hs-CRP through SES and BMI *for right-handed participants* was estimated as positive and statistically non-zero, *β* = 0.117, SE = 0.033, 95% CI (0.053 - 0.181). For left-handed participants, this effect was estimated as no different from zero, *β* = 0.152, SE = 0.112, 95% CI (-0.067 - 0.372). All other indirect effects specified were estimated to be statistically no different from zero, however, the view is taken that without other evidence, it is plausible that the focal indirect effect estimated for left-handed participants is estimated as no different than zero due to sampling error rather than grounds to assert differences in the inflamamtory cascade, so hypothesized here, because of neuroanatomical/functional group differences.

**Table 6.** Indirect Effects.

| Table 6. Indirect Effects |  |  |  |  |
| --- | --- | --- | --- | --- |
| Parameter | Estimate | Std. Error | Lower | Upper |
| Indirect (Serial): Discrimination → SES → BMI → CRP (Right-Handers) | 0.117 | 0.033 | 0.053 | 0.181 |
| Indirect (Serial): Discrimination → SES → BMI → CRP (Left-Handers) | 0.152 | 0.112 | -0.067 | 0.372 |
| Indirect (Serial): Discrimination → PosPsyCap → BMI → CRP (Right-Handers) | -0.005 | 0.005 | -0.015 | 0.006 |
| Indirect (Serial): Discrimination → PosPsyCap → BMI → CRP (Left-Handers) | -0.008 | 0.028 | -0.064 | 0.047 |
| Indirect: Discrimination → BMI → CRP (Right-Handers) | 0.014 | 0.066 | -0.115 | 0.143 |
| Indirect: Discrimination → BMI → CRP (Left-Handers) | -0.066 | 0.194 | -0.445 | 0.314 |
| Difference (Direct): Discrimination → SES (Right vs. Left) | -0.054 | 0.136 | -0.320 | 0.212 |
| Difference (Serial Indirect): via SES (Right vs. Left) | -0.036 | 0.115 | -0.261 | 0.190 |
| Difference (Serial Indirect): via PosPsyCap (Right vs. Left) | 0.004 | 0.029 | -0.053 | 0.061 |
| Total Effect: Discrimination → CRP (Right-Handers) | 0.172 | 0.271 | -0.359 | 0.702 |
| Total Effect: Discrimination → CRP (Left-Handers) | -0.168 | 0.882 | -1.897 | 1.560 |
| Difference (Total Effect): Discrimination → CRP (Right vs. Left) | 0.340 | 0.919 | -1.461 | 2.142 |
| Note: Only the estimate of the serial, (partial) mediation effect for *right-handers of Discrimination through SES through BMI on the inflammatory marker (C-Reactive Protein) is statistically non-zero. |  |  |  |  |

**Figure 4.**
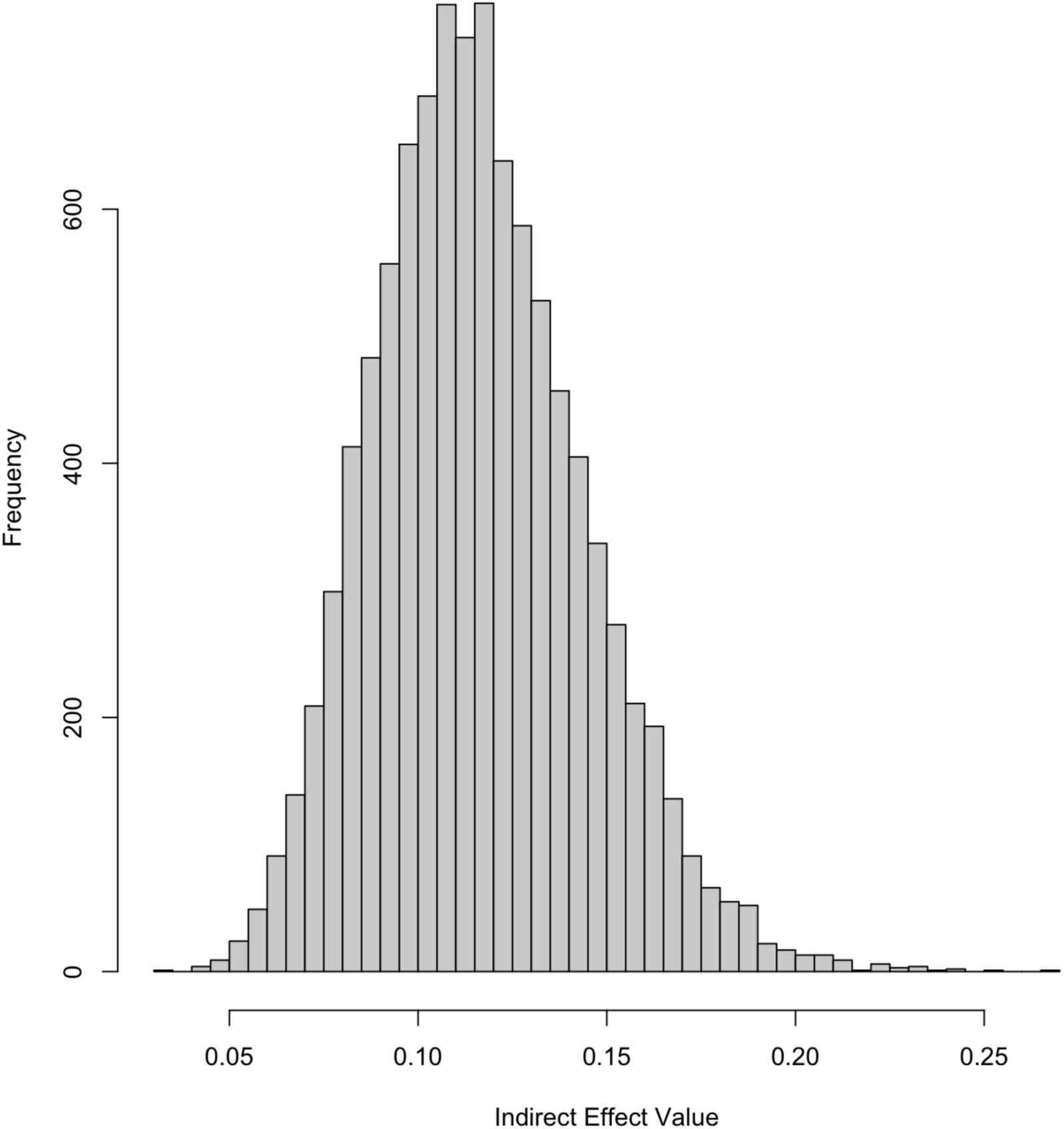
Bootstrap Distribution for ind_SES_BMI_g1

#### Bayesian Estimates

Posterior distributions and 95% credible intervals for the specific indirect effect of Discrimination on CRP through Socioeconomic Status and then Body Mass Index (on the inflammatory marker, hs-CRP) support the MLR point estimates and confidence intervals. Moreover, the standardized Bayesian estimates (Figure 5 and Table 7, above) provide comparability of model weights. While this pathway is estimated non-zero by both models, a sense of the comparability of these weights (influencing our manifest proxy for systemic inflammation) is provided by the Bayes estimates (with credible intervals) as well, where the standardized effects of BMI, sex, and the indirect effects are displayed.

**Figure 5.**
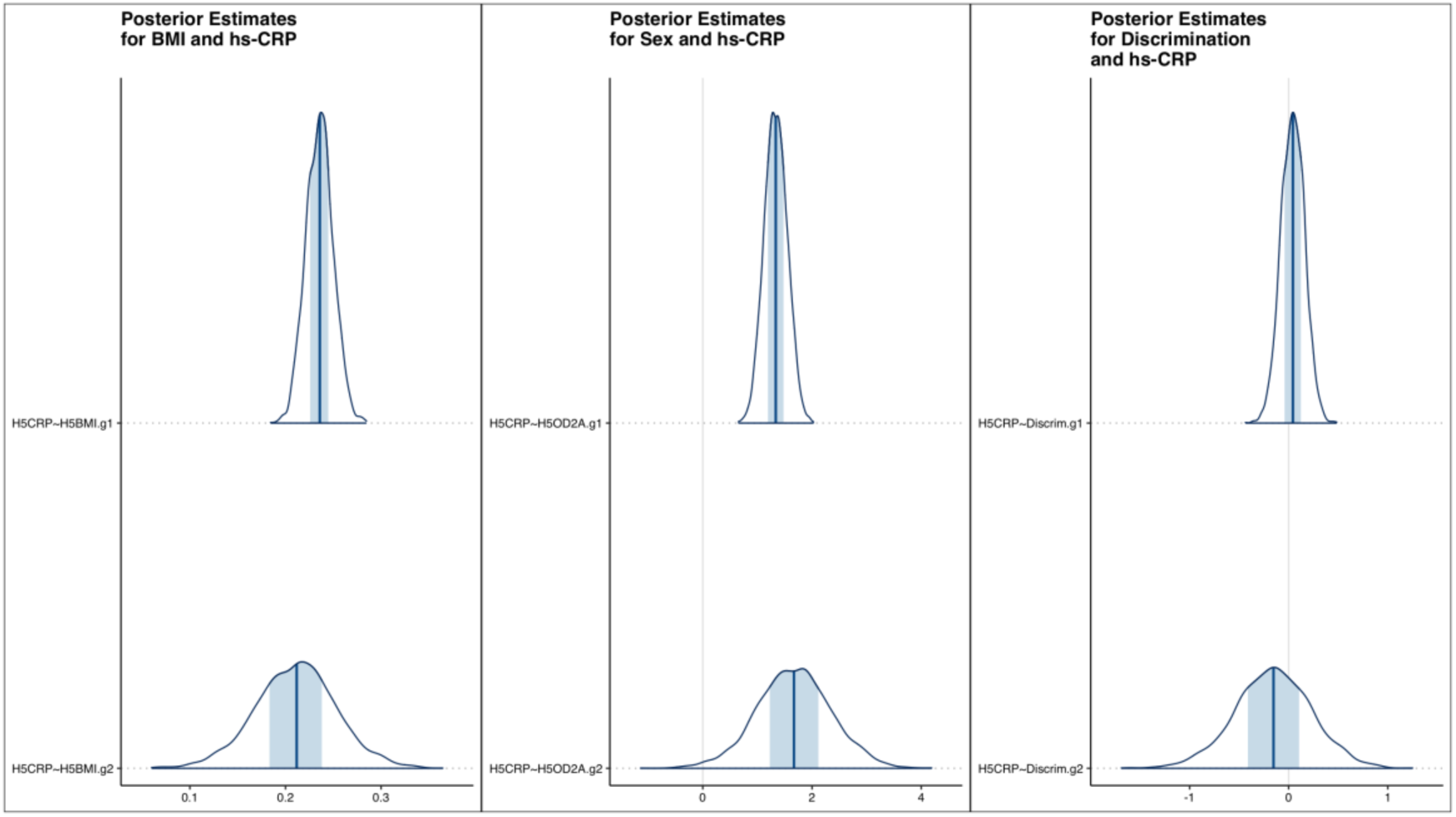
Bayesian Parameter Estimates

**Table 7.** Followup Estimates (MCMC)

| | Outcome | op | Predictor | Group | $\beta$ | SE | Lower | Est | Upper |
| --- | --- | --- | --- | --- | --- | --- | --- | --- | --- |
| h1_g1 | H5CRP | ~ | H5BMI | 1.00 | 0.44 | 0.01 | 0.21 | 0.24 | 0.27 |
| h1_g2 | H5CRP | ~ | H5BMI | 2.00 | 0.44 | 0.04 | 0.13 | 0.21 | 0.29 |
| sexCRP_g2 | H5CRP | ~ | H5OD2A | 2.00 | 0.20 | 0.67 | 0.35 | 1.66 | 2.98 |
| sexCRP_g1 | H5CRP | ~ | H5OD2A | 1.00 | 0.17 | 0.21 | 0.94 | 1.34 | 1.74 |
| ind_SES_BMI_g1 | ind_SES_BMI_g1 | := | d_g1*g1_g1*h1_g1 | 0.00 | 0.03 | 0.02 | 0.07 | 0.12 | 0.16 |

The MLR estimates are supported by the Bayes estimates. Above, Group 1 (g1) indicates estimates for right-handed participants. All parameters estimated with Rhat >= 0.999

The indirect effect (influencing the focal outcome, hs-CRP) involves the synergistic influence of discrimination, socioeconomic status (SES), and BMI. As displayed above (Figure 6), with increased SES, CRP is predicted to be reduced (relative to low SES participants) in each context of perceived discrimination - except for those around 1 SD lower than average for BMI among those who experience the most discrimination. For these participants, the trend reverses, with those lowest in SES actually predicted to have lower inflammatory markers than those at the highest end of SES (as sampled). Additionally, a Johnson-Neymon plot of the indirect effect is provided in Figure 7, illustrating the estimated range of perceived discrimination and SES where a non-zero influence to CRP is expected, given the sample.

**Figure 6.**
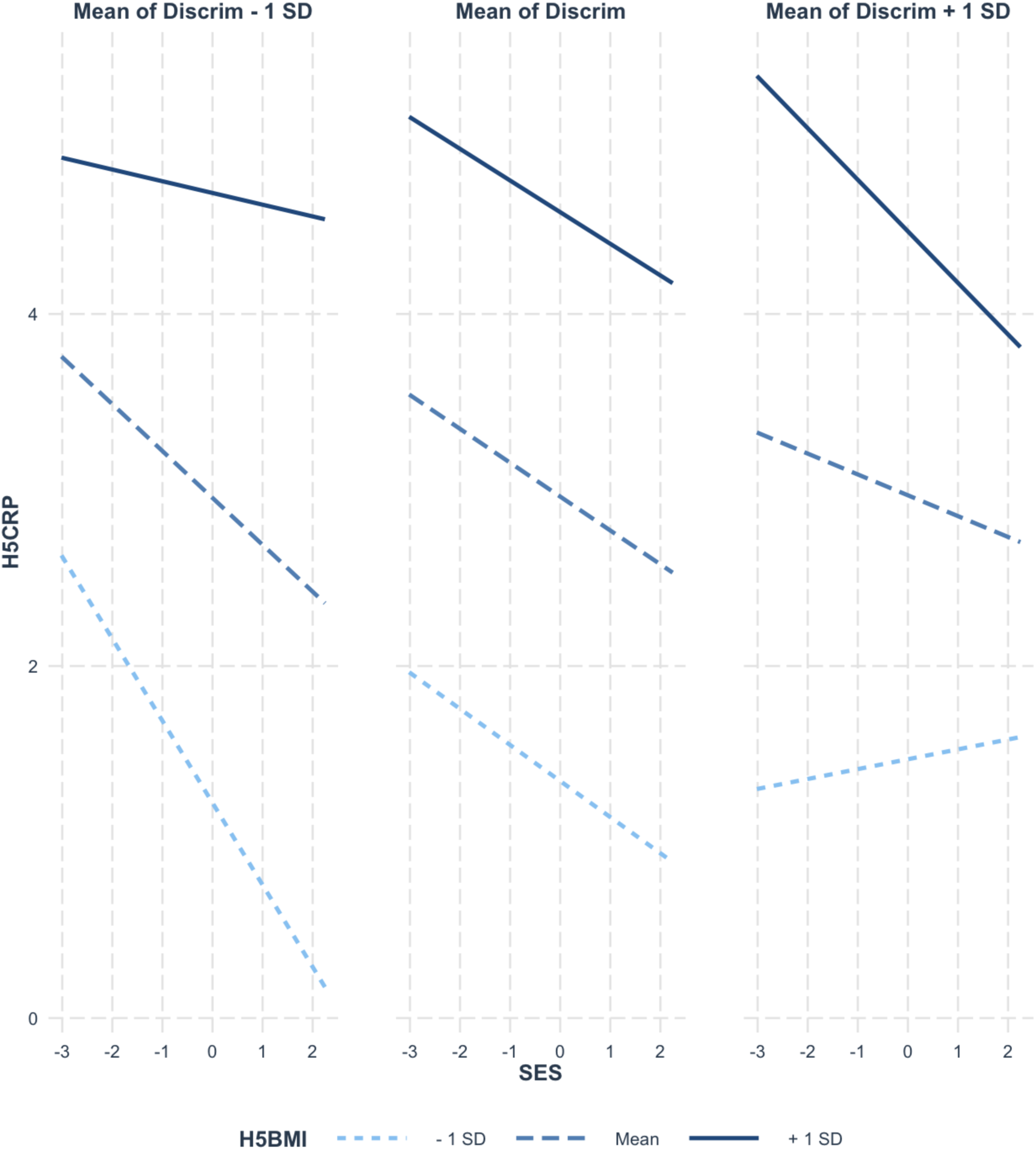
Estimated Contextual Effects

**Figure 7.**
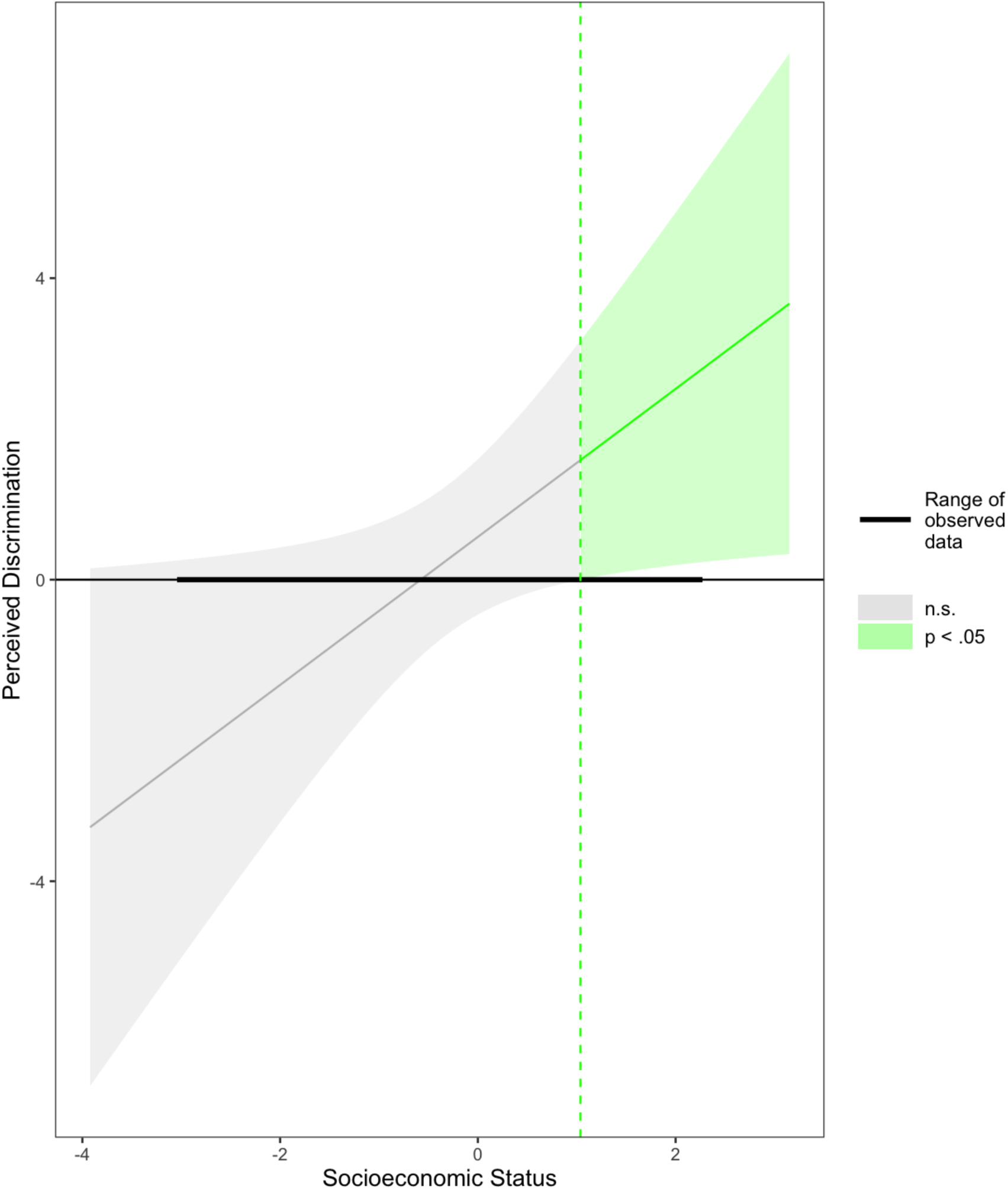
The Slope of Discrimination on CRP

## Discussion

This conceptualization of risk (inflammatory marker outcome) tests a suite of direct and indirect effects, estimated in parallel using an ADD Health-based (wave five) sample. BMI’s positive association with hs-CRP stands as the strongest contributor to variance in the inflammatory outcome; next are the estimated effects of sex (both groups). Inflammatory markers are routinely associated with adiposity and obesity to the extent that some researchers refer to groups of common measures as obesity-related inflammatory markers (ORIM); this includes C-Reactive Protein (Forsythe, Wallace, and Livingstone, 2008). Forsythe et al. (2008) showed improvements in ORIMs (declines) appeared to coincide with weight loss in participants among studies reviewed. Ferrante (2007) argues that obesity increases the frequency and changes the status of certain organelles in adipose tissue and that this accounts for some of the inflammation that may arise with obesity and many individuals relatively higher on the BMI scale.

For the analysis sample, there isn’t support for an influence on the inflammatory marker (hs-CRP) from perceived discrimination when BMI is included; while an indirect path is retained-it’s influence on systemic inflammation isn’t as pronounced as the direct effect of BMI or sex differences; the indirect effect estimate does allow us to convey predicted effects across contexts. In this case, in all but one context, a protective effect (lower crp) is predicted to coincide with higher SES at sample mean BMI, a standard deviation below (BMI), and a standard deviation above (‘pick-a-point’). Previous work already established concern for the effects of discrimination on inflammation and downstream health effects, raising susceptibility to cardiovascular disease, poorer oral health, and metabolic syndrome. From this, a question stands: what can/should be done to minimize the observed detrimental effects? BMI may stand as a modifiable (and replicated) target, bolstering efforts for increased/improved physical activity and improved nutritional knowledge/access as means to not only improve the overall health of people, but also as a facet of resilience that can be strengthened. Sensible considerations can be made inclusive of the important constructs estimated. Lower SES may be a primary cause of fewer opportunities and resources conducive to healthy lifestyles, which, in turn, may influence hs-CRP and inflammation for many people. While aims to prevent undue inflammation and associated disease states may benefit from a focus on working towards healthier body composition (and BMI), analyses and interventions that account for a more nuanced cascade of interconnected psychosocial constructs (such as discrimination, psychological capital, etc.) may be tenable and necessary to address health disparities. While the indirect effect was not observed for the left-handed group of participants, the view is taken that statistical power may be the reason for this (considerably fewer participants in this group) rather than an assertion of an empirical regularity wherein left-handed participants, would be expected to be somehow invulnerable to psychosocial health impacts. This underscores the importance of sample size for detecting small, statistically significant effects and here, to observe effects relevant for a participant group that may be less well-understood. Since this group was previously observed to be associated with atypical brain-immune interactions (Stoyanov, Decheva, Pashalieva, and Nikolova, 2012) and rheumatoid arthritis (Yaku, Hashimoto, Furu, Ito, Yamakawa, Yamamoto, Fujii, Matsuda, Mimori, and Terao, 2016), for instance, more can be learned about the psychosocial-biological phenomena that may influence inflammation and downstream progressions of disease.

### Potential Confounds

#### Reverse Causation

While the main finding (Discrimination → SES → BMI → hs-CRP) assumes the effects of everyday discrimination may influence SES, influencing BMI and potentially causing detrimental changes in hs-CRP, it is not impossible that the converse could be true; those with relatively higher inflammation are prone to higher BMI and this results in increased perceived discrimination. The path tested, here, seems *more* biologically plausible, however, one should leave room for the possibility that the reverse could be true - at least in some conditions, groups, or ranges of these dimensions sampled.

## Data Availability

All source data are publicly available.

https://addhealth.cpc.unc.edu

## Appendix

### Data

#### Linear Regression

##### Crossover Interaction: BMI and Handedness

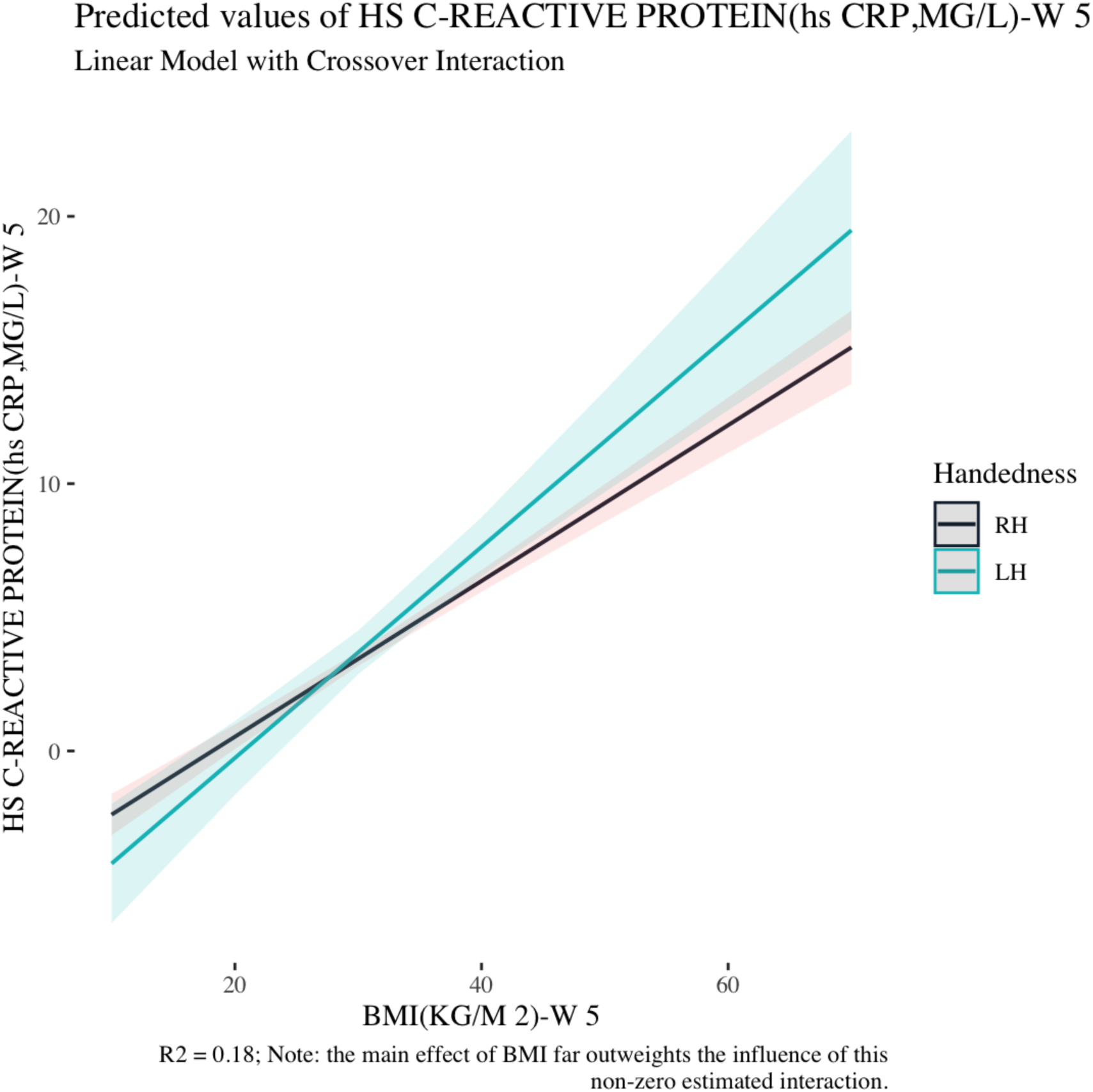

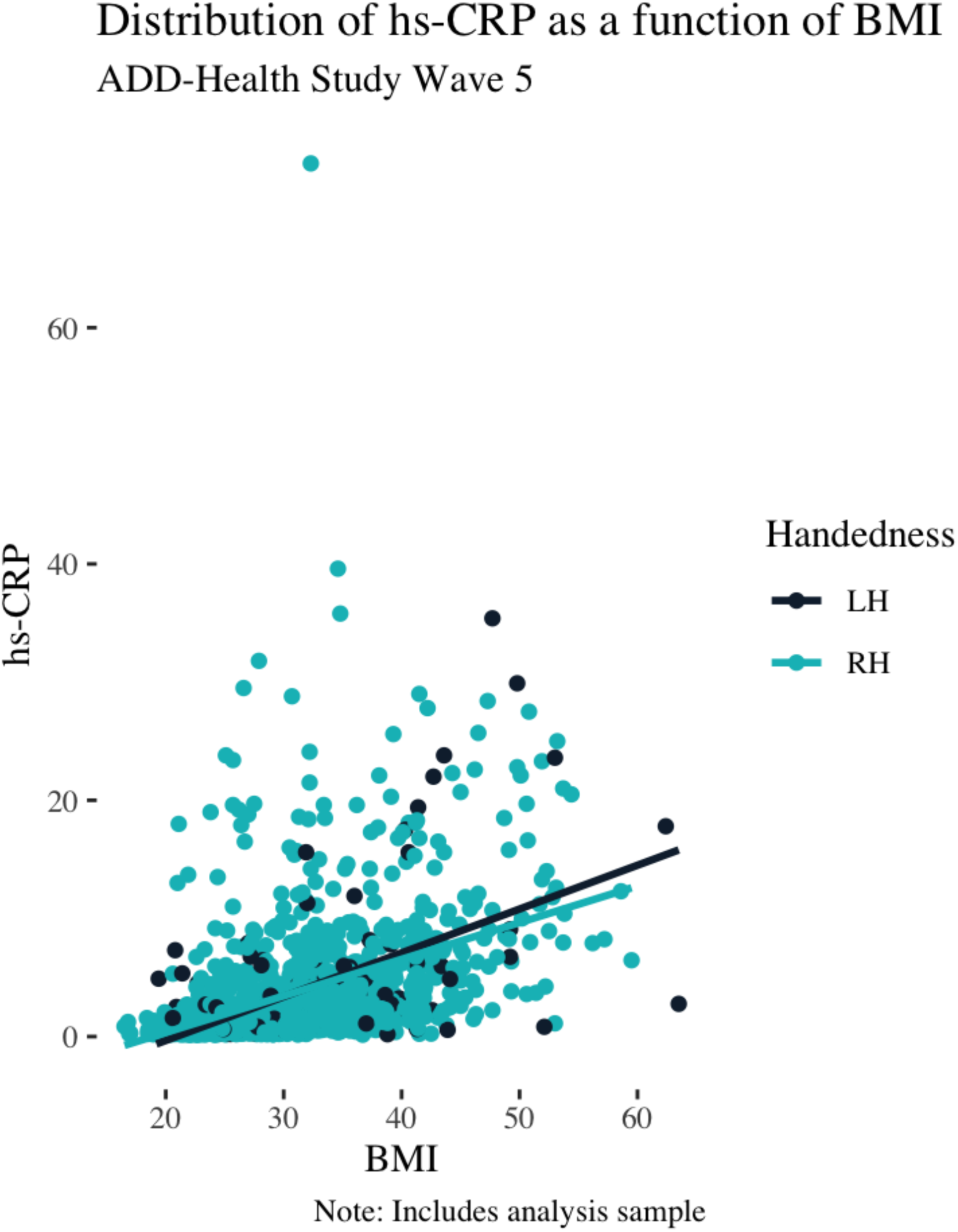

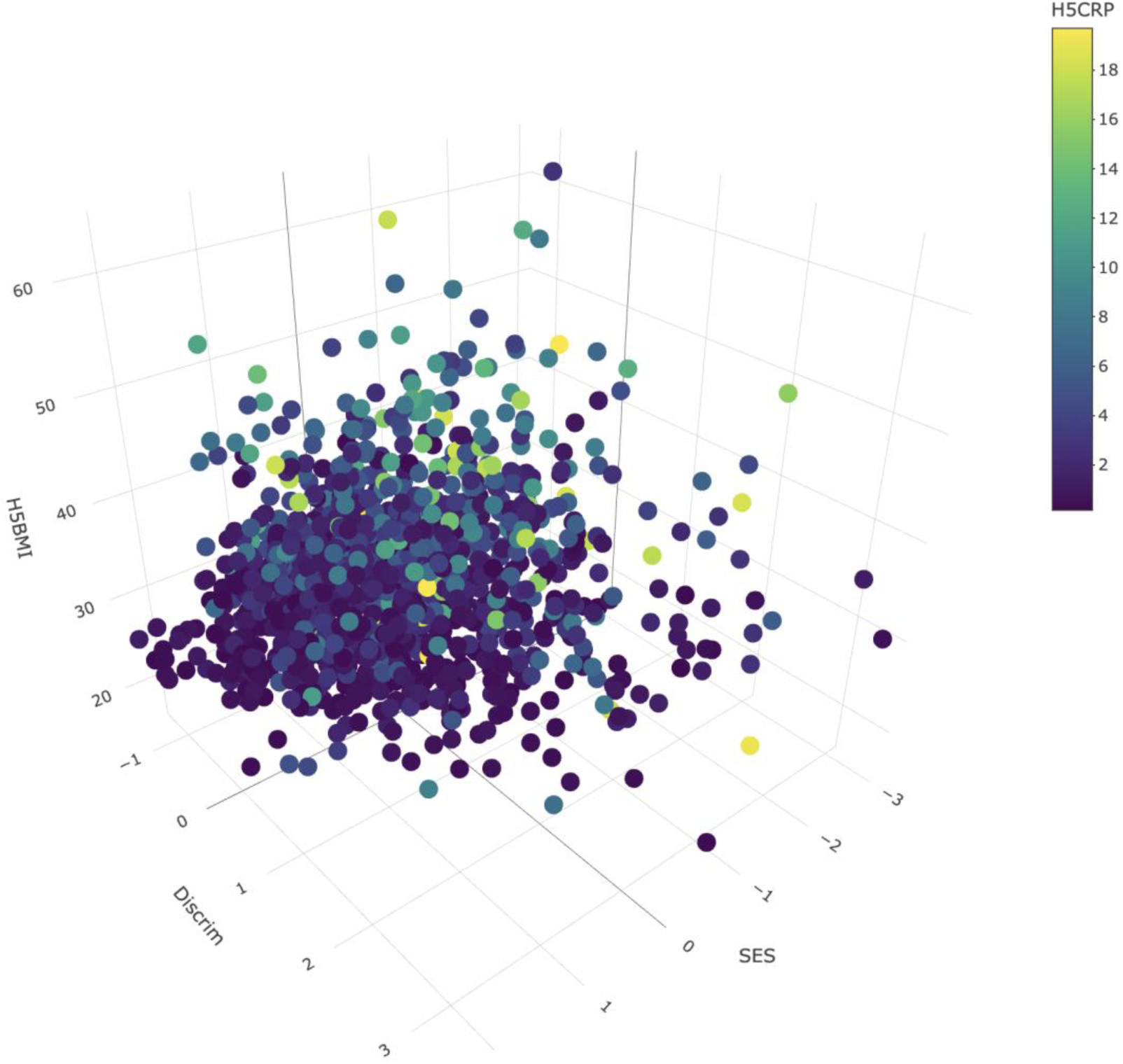

Above, the predicted factor scores are projected with the manifest variables: BMI and CRP (the latter being the ORIM outcome)

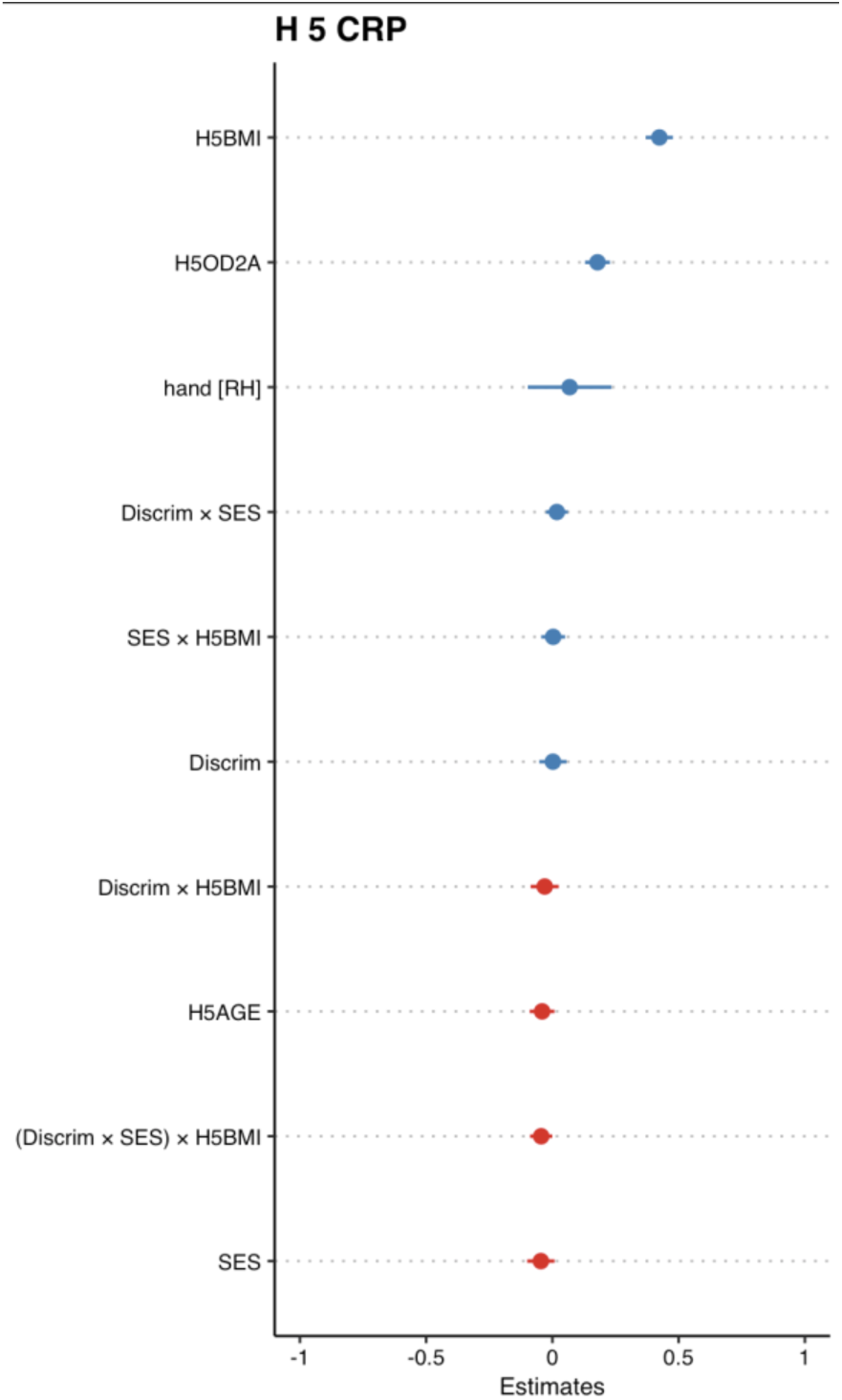

The relative magnitude of the estimated effects are shown above. Here, positive effects are shown in blue and negative estimates in red. Effect sizes were calculated for direct effects, suggesting the strongest direct effects were associated with BMI, sex, and SES, *η_p_*^2^ = 0.18, *η_p_*^2^ = 0.041, and *η_p_*^2^ = 0.040 (note: in the plot above, standardized weights are shown rather than partial eta squared, the proportion of variance in the dependent variable that is attributed to an independent variable, while controlling for the effects of other variables in the model).

